# Development of a cost-effective multiplex quantitative RT-PCR assay for early detection and surveillance of Dengue, Chikungunya, and co-infections from clinical samples in low-resource settings

**DOI:** 10.1101/2024.09.10.24313257

**Authors:** Shruthi Uppoor, Samruddhi Walaskar, Ritika Majji, SP Deepanraj, K.V Thrilok Chandra, H.N Madhusudan, A.S Balasundar, Rakesh Kumar Mishra, Farah Ishtiaq, Mansi Rajendra Malik

## Abstract

**Background:** Dengue and Chikungunya are Aedes-borne diseases that are predominantly prevalent in tropical and subtropical regions, affecting public health globally. Dengue is caused by multiple antigenically different Dengue virus (DENV) serotypes (DENV-1 to DENV 4) in the Flaviviridae family and Chikungunya (CHIKV) in the Togaviridae family. Both viral diseases produce similar clinical manifestations, especially in the early stages of infection which poses a significant challenge for timely diagnosis and improper disease management. In India, diagnosis of Dengue and Chikungunya relies on ELISA-based tests, which often lead to false negatives and under estimation of the disease burden.

**Methods:** A multiplex, quantitative, real-time PCR assay, DENCHIK was developed for simultaneous detection of DENV serotypes and CHIKV.A total of 903 sera samples were screened from suspected febrile patients across 161 public health centers in Bengaluru, between July 2022 - December 2022. The sensitivity and specificity of DENCHIK assay was compared with ELISA (NS1 antigen and Immunoglobulin M (IgM) antibodies) and two commercially available q RT-PCR assays for DENV and CHIKV.

**Findings:** Using DENCHIK assay,36% infections were DENV, 17% CHIKV and 8% were DENV CHIKV co-infections. In contrast, ELISA detected 29.90% of DENV and 22.92% of CHIKV infections. We observed 9% prevalence of DENV infections using NS1 ELISA as compared to 24% by IgM ELISA. DENV-1 was the predominant serotype followed by DENV-2, DENV-3 and DENV-4. There was an increase in the prevalence of DENV and CHIKV infections from June to September 2022, coinciding with the monsoon season. There was no significant difference observed in the prevalence of DENV and CHIKV infections across genders and ages. The sensitivity and specificity of DENCHIK assay in DENV detection as compared to NS1 ELISA assay was observed to be 62.82% and 66.45%, respectively. In comparison to commercially available q RT-PCR assays for DENV detection, DENCHIK assay exhibited 99% and 98% sensitivity and specificity, respectively. Similarly, in case of CHIKV 26% sensitivity, 86% specificity and 98% sensitivity and specificity were observed, as compared to the IgM ELISA and commercial RT-PCR assays, respectively.

**Conclusion:** DENCHIK assay successfully enabled, simultaneous amplification of all four DENV serotypes and Chikungunya, from clinical samples. DENCHIK assay detected 7.6% of additional Dengue infections and 6.65% less of Chikungunya infections in clinical samples, as compared to detection by ELISA. As, compared to ELISA, DENCHIK demonstrates early and accurate detection of DENV and CHIKV with higher sensitivity and specificity, as early as day one of symptom onset post infection. DENCHIK aids in estimating the exact prevalence of DENV and CHIKV infections, that are often misdiagnosed, using ELISA. Molecular surveillance using targeted diagnostic assays such as DENCHIK could be used to determine the prevalence of multiple DENV serotypes, CHIKV and DENV-CHIKV Co-infections from clinical samples. The findings from the study shall be useful to inform and aid the public health authorities, to contain and curb the rapid spread of these diseases in the community.

**Author Summary:** Dengue and Chikungunya are most common arboviral illnesses affecting more than half of the world’s population. Both the viral diseases have overlapping symptoms, which poses a challenge for accurate differential diagnostics in low-resource setting. Infection with one or more different serotypes of DENV results in a phenomenon, known as antibody-dependent enhancement (ADE), wherein antibodies against one serotype, instead of protecting against DENV infection caused by other serotypes, aids in the viral uptake by the host immune cells, resulting in severe dengue.

Rapid antigen tests targeting NS1, and IgG/IgM are the most common methods used to detect DENV and CHIKV infections. However, there are several limitations of serological assays: a) ELISA cannot differentiate DENV serotypes, b) depending on the stage of infection, ELISA-based tests often provide false-positives or false-negatives. This warrants a need for a reliable molecular method which can differentiate between DENV serotypes and across Dengue and Chikungunya with reasonable sensitivity and specificity.

Bengaluru has highest dengue burden in Southern India. There is high infestation of *Aedes aegypti* and *Aedes albopictus* in diverse breeding habitat and year-round circulation of four serotypes. Currently, Dengue and Chikungunya testing relies on ELISA (NS1, IgM and IgG) often leading to under estimation of disease burden. To address this gap, a cost-effective multiplex qRT-PCR assay, DENCHIK was developed for simultaneous detection of four DENV serotypes and CHIKV. The sensitivity and specificity of DENCHIK assay was tested across months and days from onset of febrile symptoms and compared with ELISA and two commercially available kits. We suggest implementation of molecular methods and using DENCHIK assay in urban health centres would help reduce underestimation of cases, actual estimates of disease burden across seasons and help in better clinical management of Dengue and Chikungunya.

## 1. Introduction

Dengue virus (DENV) and Chikungunya Virus (CHIKV) are arboviruses, transmitted by *Aedes aegypti* and *Aedes albopictus* mosquitoes across the tropical and sub-tropical regions. DENV is a single stranded positive sense RNA virus and belongs to the family Flaviviridae [1]. It consists of four antigenically distinct, however 65-70% homologous serotypes known as DENV-1, DENV-2, DENV-3 & DENV-4 [2]. CHIKV is an enveloped virus that belongs to the family *Togaviridae* and the genus *Alphavirus*. DENV and CHIKV infections are endemic in Asian and African subcontinents with year-round transmission[3]. Both arboviruses have seasonal patterns of transmissions with a peak before the onset of monsoon season[4]. Urbanisation, human-made habitat, humidity, temperature are some of the factors contributing to mosquito densities and disease dynamics. However, recent expansion in the geographical ranges of two *Aedes* species, in Europe and the Americas has led to the emergence of locally transmitted dengue cases [1,5,6].

Both DENV and CHIKV incubation period ranges from 2 days until 10 days. Acute infection can lead to mild undifferentiated acute febrile illnesses, making it clinically indistinguishable from other viral, bacterial, and parasitic infectious diseases such as influenza, chikungunya, leptospirosis, filaria, and malaria [7–9]. Co-circulation of DENV and CHIKV in the same region and overlapping symptoms and common clinical presentations make the differential and accurate diagnosis of DENV and CHIKV challenging. For instance, DENV infection can lead to dengue shock and haemorrhagic fever, while CHIKV infection has been associated with a persistent arthralgia for months after the infection has subside[10]. Misdiagnosis often lead to false alarms, underestimation of cases and clinical management of disease[11].

Early diagnosis of Dengue remains challenging as DENV viremia is detectable within 24-48 hours before the onset of symptoms and continues for 5-6 days allowing a short window to detect the NS1 protein in blood/serum[12]. In India, routine diagnosis of DENV and CHIKV is based on serological methods for non-structural Protein 1 (NS1) antigen and Immunoglobulin M (IgM) and Immunoglobin G (IgG) levels, respectively. Patients suspected of Dengue fever after 6 days are usually tested using IgM and IgG antibodies [13]. Furthermore, cyclic DENV outbreaks in the endemic regions occur every 2–4 years, often associated with serotype/genotype replacement, where serotype/genotype dominance changes during the subsequent outbreak [14]. Detection of DENV serotypes has utmost clinical and epidemiological importance to understand disease severity and which cannot be ascertained using currently used serological methods. Intermediate level of cross-reacting antibodies in patients with previous DENV infection, and the presence of heterotypic serotypes in infected patients might increase the severity of the disease also known as the Antibody dependent enhancement (ADE)[15]. Hence, the detection of mixed serotype infection could be a very useful resource for the clinicians to timely prioritize treatment of DENV infected patients.

Bengaluru has highest dengue burden in Southern India. There is high infestation of *Aedes aegypti* and *Aedes albopictus* in diverse breeding habitat [16] and year-round circulation of four serotypes. Currently, Dengue and Chikungunya testing relies on ELISA (NS1, IgM and IgG) often leading to under estimation of disease burden. To address this gap, we designed an in-house, cost-effective, TaqMan probe-based, multiplex (five-plex), quantitative reverse transcriptase PCR (qRT-PCR) assay, to detect and quantify all four DENV serotypes and Chikungunya simultaneously. We conducted a clinical surveillance study to understand the variance in the detection of DENV and CHIKV infections using routine serology (ELISA) and nucleic acid-based amplification (multiplex q RT-PCR) on sera samples from suspected febrile patients in Bengaluru city. In addition, we propose a systematic diagnostic approach based on the days post onset of symptoms and implementation of molecular testing to enhance early detection of DENV and CHIKV for timely treatment and management of these diseases.

## 2. Methods

### 2.1 Ethics Statement

The study was approved by the Institutional Human Ethics Committee, Institute for Stem Cell and Regenerative Medicine, Department of Biotechnology, Government of India (Reference number: inStem/IEC-26/04N) and Ethics Committee of Bangalore Medical College and Research Institute, Bangalore, Karnataka (Reference no: BMCRI/PS/41/2022-23).

Informed consent was obtained from all patients enrolled in the study, as per ICMR guidelines. The study procedure, description and questionnaire were explained verbally and provided as a document both in English and the local language (Kannada) for adult patients. For adolescents and young children, parental approval and consent from the study participants were obtained verbally and in writing.

### 2.2 Study design

We conducted a six-month longitudinal study between July 2022 and December 2022 in urban Bengaluru in collaboration with the local civic body of Bengaluru, the Bruhat Bengaluru Mahanagara Palike (BBMP). A total of 903 blood samples were collected from suspected febrile patients as per WHO inclusion criteria (18) (e.g., high fever (>37.8°C), myalgia, headache, retroorbital pain, arthralgia, and gastrointestinal disorders) (Supplementary Table 1). Blood samples were collected from 161 collection points comprising of the Urban Primary Health centres, Referral hospitals and maternity homes under the aegis of BBMP. In addition, metadata such as the days post onset of symptoms, and the nature of symptoms, age and gender was recorded.

Blood samples were processed for sera and tested for DENV and CHIKV using ELISA (NS1 and IgM antibodies) [11,16] at Hitech Labs at the H. Siddaiah Road Referral Hospital in Bengaluru by Dr Shruthi Uppoor (SU). Subsequently, the remaining serum was tested using in-house multiplex qRT-PCR for molecular detection at the Tata Institute for Genetics and Society by Dr Mansi Rajendra Malik (MRM) (Fig 1).

**Fig. 1:**
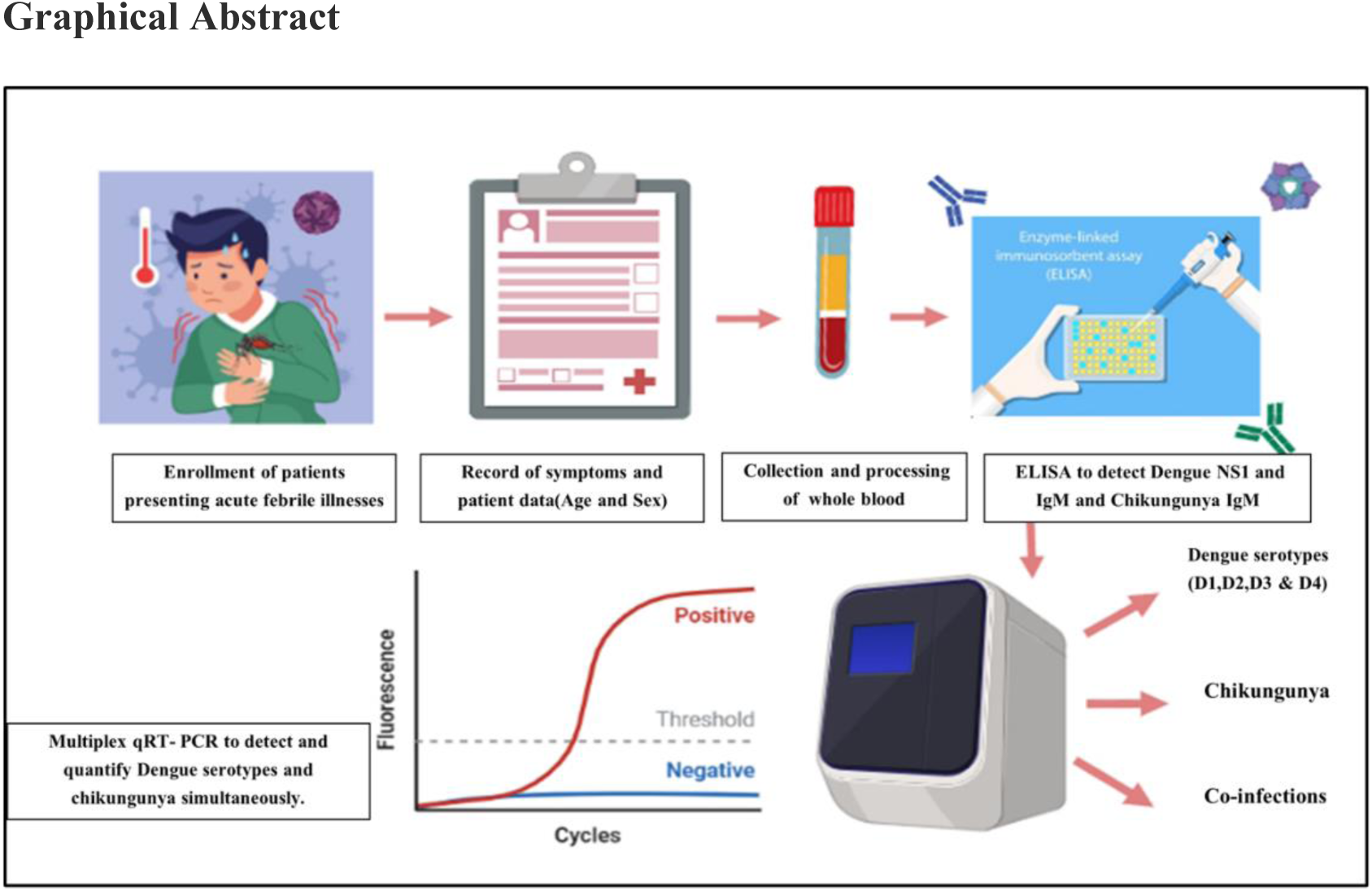
Workflow of molecular surveillance of DENV and CHIKV from sera of suspected febrile patients.

### 2.3 Serological testing of DENV and CHIKV

The detection of DENV NS1 antigen and IgM antibodies from sera samples was conducted using a one-step, sandwich ELISA developed by the Pan bio–Dengue Early (Pan Bio-Diagnostics, Brisbane, Australia) and the IgM-capture ELISA kit (Pan Bio-Diagnostics, Brisbane, Australia) [17,18]. For CHIKV, IgM Capture ELISA Kit developed by National Institute of Virology (Arbovirus Diagnostic NIV, Pune, India) was used as per manufacturer’s instructions [19–21].

### 2.4 Development of molecular assay for DENV Serotypes (1-4) and CHIKV (DENCHIK assay)

#### 2.4.1 Viral RNA extraction

Viral RNA was extracted from 140 µl serum samples using QIAamp Viral RNA kit (QIAGEN, Hilden, Germany) as per the manufacturer’s instructions. The extracted viral RNA was quantified spectrophotometrically using The Nanodrop 2000 (ThermoFisher Scientific, USA) [16,22]. For q RT-PCRs, 25 ng of the extracted viral RNA was used.

#### 2.4.2 Primer Design

For designing DENV serotypes and CHIKV primers, following DENV sequences from GenBank (NCBI: DENV-1 (NC_001477), DENV-2 (NC_001474.2), DENV-3 (NC_001475.2), DENV-4 (NC_002640.1), CHIKV (MK370030.1). After checking for ambiguities in the sequences, the most conserved gene sequences across the four serotypes were selected for primer design. Primers covering the Polyprotein genes (poly) were chosen for DENV-1, DENV-2, and DENV-3, while the E gene was selected for DENV-2 and CHIKV.

The primers and probes were designed inhouse and their specificity to respective Dengue serotypes and CHIKV was assessed using NCBI BLAST (Table 1a and Table1b).

**Table 1a).**
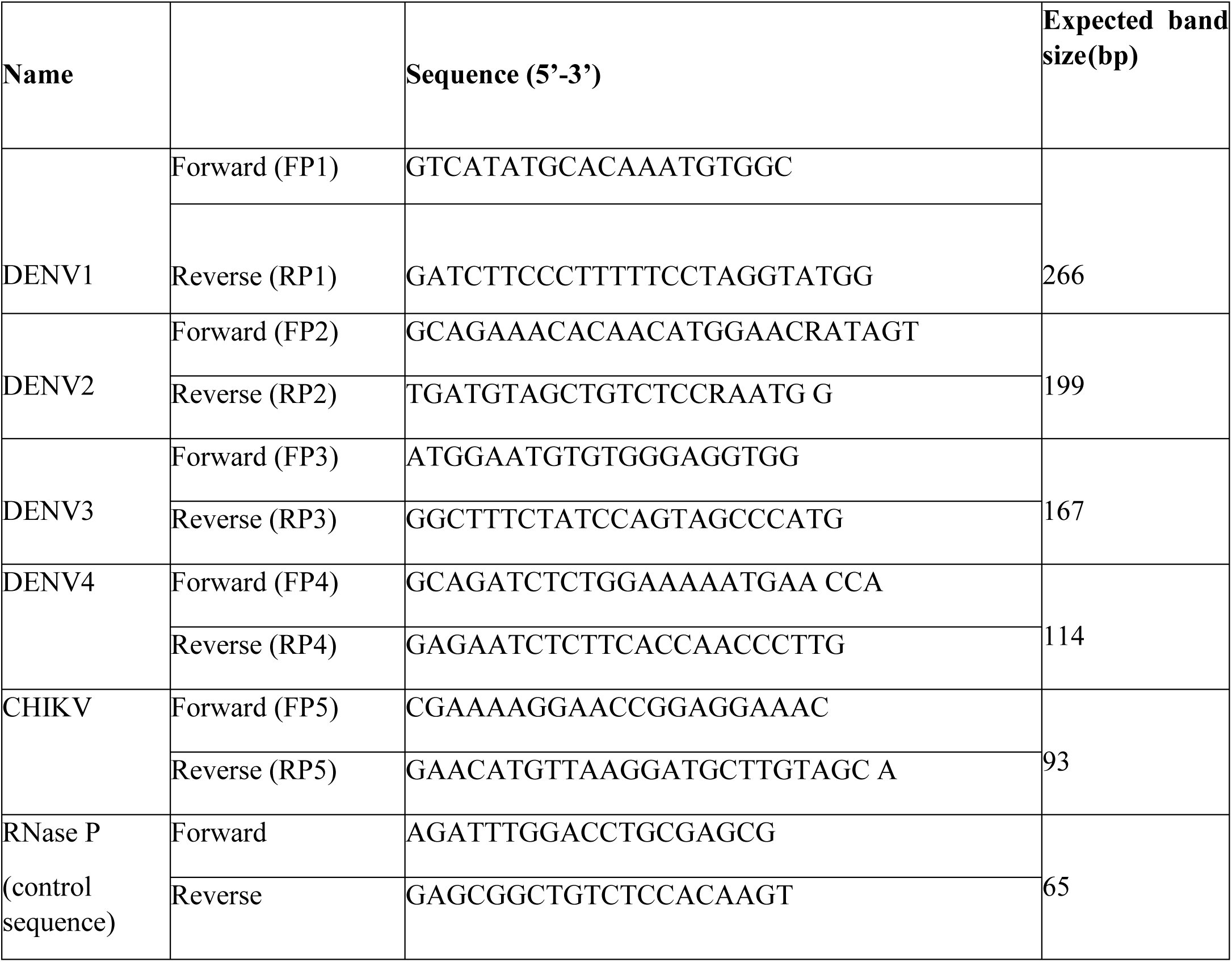
Primer sequences for DENV serotypes, CHIKV, and RNase P gene as internal control.

**Table 1b:**
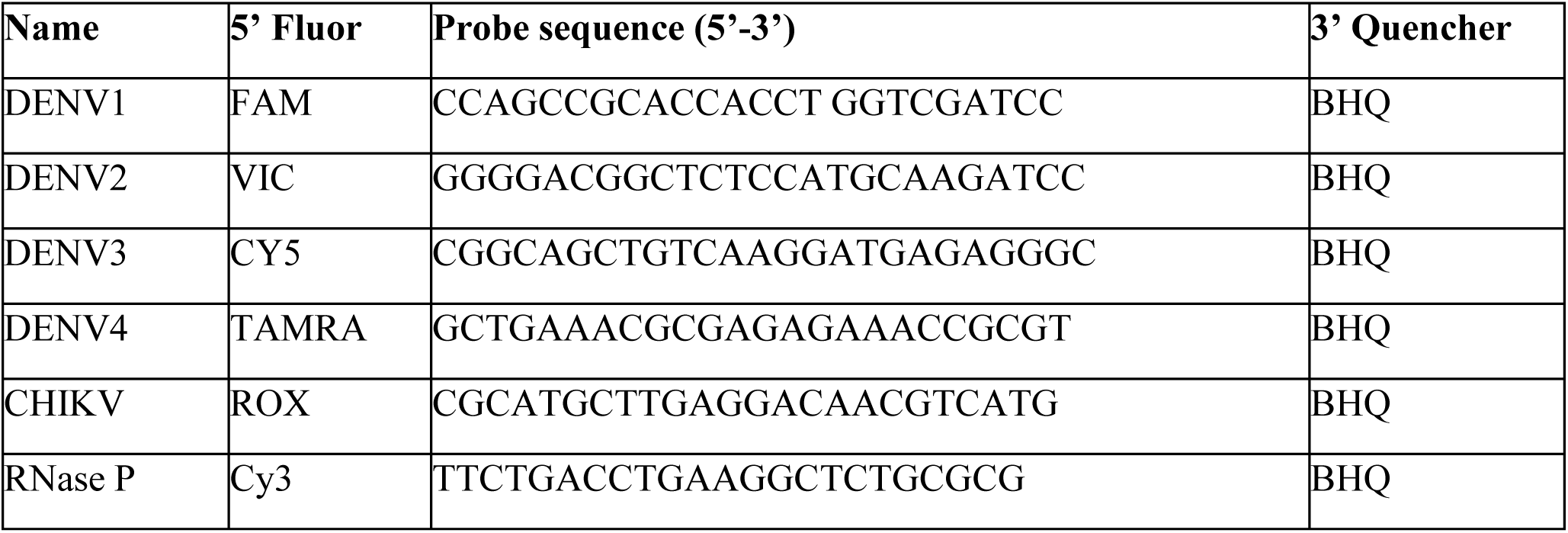
Probe sequences for DENV serotypes, CHIKV, and Internal control (RNase P).

#### 2.4.3 Quantification of DENCHIK assay

The respective DNA sequences for DENV serotypes and DENV encompassing the designed primer and probe sequences were synthesized and cloned into the plasmid vector pBluescript II KS (+), provided by GenScript, USA. The plasmids were then linearized via PCR using the forward primer GTAACGCCAGGGTTTTCCCAGTC and the reverse primer CACAGGAAACAGCTATGACCATGATTAC to generate the DNA amplicons for subsequent invitro transcription (Supplementary Table 2). The primer pairs were used with Q5 high fidelity DNA polymerase (NEB, USA) for PCR using the following parameter conditions: 98°C for 5 min, 98°C for 30 s, 68°C for 30 s, and 72°C for 45 s for 28 cycles, and finally at 72°C for 5 min. The PCR was performed on a C1000 Touch thermocycler (Bio-Rad, CA, USA) and the products were run on a 1% agarose gel. The respective Dengue serotypes and Chikungunya amplicons were excised, and gel extraction and purification were performed using the Monarch DNA Gel extraction kit (NEB, USA) and Monarch PCR clean-up kit (NEB, USA), respectively.

#### 2.4.4 In vitro transcription and RNA purification

Ten microliters of purified and linearized Dengue serotypes and Chikungunya amplicons were used for T7 polymerase-mediated *invitro* transcription using the Hiscribe T7 high-yield RNA synthesis kit (NEB, USA) [22,23]. The reaction was performed according to the manufacturer’s instructions and the resulting RNA transcripts were purified using phenol chloroform purification method [24]. The concentration of the purified RNA transcripts was measured (ng/μl) using a Nanodrop 1000 (Thermo Fisher). The specificity of DENCHIK assay was compared with closely related flaviviruses and the assay was found to be highly specific for DENV serotypes and CHIKV with no cross reactivity with synthetic gene fragments of Hepatitis A virus, Hepatitis C virus, Hepatitis E virus, Japanese encephalitis virus, Zika virus and Kyasanur Forest disease virus. Further, RNA from 50 serum samples from healthy individuals were tested with primers and probes used in DENCHIK assay and no amplification was observed [16,25].

#### 2.4.5 Performance evaluation of the DENCHIK assay

The purified RNA transcripts were assayed in the multiplex q RT-PCR assay in triplicates of ten-fold serial dilutions, ranging from 10^9^ to 1copy(s)/microlitre. The Limit of Detection (LoD) was determined by performing the qRT-PCR assays using the RNA dilutions. The standard curve was generated by plotting the CT values with the RNA dilutions. The copy number per microlitre was calculated for each DENV serotype and CHIKV using the following equation [22,25] (Supplementary Table 3).

RNA molecules per microlitre= [(g/µl)/ (transcript length in nucleotides X 340)] X 6.022 × 10^23^.

### 2.5 Screening of clinical samples

For screening and detection, we set a 10 μl PCR reaction comprising of 2.5 µl of Luna® Probe One-Step RT-qPCR 4X Mix with UDG (#M3029E, NEB Biolabs, Ipswich, MA, USA), 50 ng/µl of RNA template, 300 nM of primers, and 250 nM of probes for each DENV (1-4) and CHIKV specific forward and reverse primers and probes (Table 2). Human RNase P was used as an internal control, and amplification was observed for all samples in triplicate. Thermal cycling parameters of this assay included a 30 s incubation at 25°C to prevent carryover contamination, followed by a 10 min reverse transcription step at 55°C. Initial denaturation was performed at 95°C for 1 min, followed by 40 cycles of PCR at 95°C denaturation for 10 s; 54°C of annealing and extension for 45 s. RT-PCR was performed and analysed using the Quantstudio 5 (Applied Biosystems, Carlsbad, CA) [26,27].

**Table 2:** Prevalence of Dengue serotypes, Chikungunya and Dengue-Chikungunya co-infections across age, sex, and symptoms.

|  | Dengue Serotypes |  |  |  |  | Chikungunya | Dengue -chikungunya<br>Co-infections |
| --- | --- | --- | --- | --- | --- | --- | --- |
| Characteristics | D1<br>n(%) | D2<br>n(%) | D3<br>n(%) | D4<br>n(%) | Mixed<br>Serotype | n(%) | n(%) |
| AGE |  |  |  |  |  |  |  |
| 0-10 | 19(5.67) | 12(3.58) | 3(0.89) | 6(1.79) | 7(2.08) | 18(10.16) | 7(8.97) |
| 11-20 | 22(6.56) | 14(4.17) | 8(2.38) | 5(1.49) | 23(6.86) | 34(19.20) | 17(21.79) |
| 20-30 | 33(9.85) | 21(6.26) | 9(2.68) | 8(2.38) | 28(8.35) | 65(36.72) | 28(35.89) |
| 31-40 | 18(5.37) | 8(2.38) | 2(0.59) | 6(1.79) | 16(4.77) | 31(17.51) | 15(19.23) |
| 41-50 | 6(1.79) | 3(0.89) | 1(0.29) | 2(0.59) | 8(2.38) | 16(9.03) | 3(3.84) |
| 51-60 | 5(1.49) | 2(0.59) | 1(0.29) | 2(0.59) | 4(1.19) | 12(6.77) | 3(3.84) |
| > 60 | 3(0.89) | 1(0.29) | 1(0.29) |  | 3(0.89) | 12(6.77) | 6(7.69) |
| SEX |  |  |  |  |  |  |  |
| Male | 52(33.98) | 21(13.75) | 15(9.8) | 13(8.4) | 57(37.25) | 86(45.58) | 39(22.03) |
| Female | 52(28.57) | 37(20.32) | 10(5.49) | 16(8.79) | 45(24.72) | 91(51.41) | 39(22.03) |
| SYMPTOMS |  |  |  |  |  |  |  |
| Fever | 89(33.08) | 48(17.84) | 22(8.17) | 10(3.71) | 80(29.73) | 135(50.18) | 66(24.53) |
| Headache | 30(29.70) | 14(13.86) | 11(10.89) | 8(7.92) | 37(36.63) | 47(46.53) | 25(24.75) |
| Body ache/myalgia | 33(26.19) | 21(16.22) | 14(11.11) | 6(4.76) | 51(40.47) | 56(44.44) | 27(21.42) |
| Rash | 2(40) | - |  |  | 3(60) | 0 | 0 |
| Cough | 0 | 0 | 2(66.66) | 0 | 1(33.33) | 2(66.66) | 2(66.66) |
| Loss of appetite | 3(50) | 2(33.33) | 0 | 0 | 3(50) |  |  |
| Nausea/vomiting | 2(13.3) | 5(38.46) |  | 1(7.69) | 3(23.07) | 3(23.07) | 2(13.3) |
| Drowsiness | 1(20) | 1(20) | 0 | 0 | 3(60) | 2(40) | 1(20) |

### 2.6 Comparison of DENCHIK with ELISA

The prevalence, specificity, and sensitivity of DENV and CHIKV detected by DENCHIK assay was compared with the NS1 antigen ELISA and IgM ELISA as reference and vice-versa. In addition, a subset of samples (n=100) was analysed by the DENCHIK assay and the detection accuracy was calculated using commercially available Altostar Dengue RT-PCR kit (Altona Diagnostics, Hamburg, Germany) and Real Star Chikungunya RT-PCR kit (Altona Diagnostics, Hamburg, Germany) respectively.

### 2.7. Statistical Analyses

The prevalence of DENV, CHIKV and DENV-CHIKV co-infections was compared and analysed by Chi-square test, using Graph Pad Prism v. 10.0, software SanDiego, California, USA. The prevalence of DENV serotypes, CHIKV, and DENV-CHIKV co-infections was compared and analysed across age and sex.

The age group of the study spanned across 11 months until 75 years of age. The mean age of the participant cohort was observed as 26.40 ± 15.07 (Table 2) years, with a sex ratio of 0.91. Diagnostic accuracy parameters such as sensitivity, specificity, positive and negative predictive values of the ELISA and q RT-PCRs assays were calculated using Chi-square and Fisher’s exact test[28]

The percent concordance and discordance (also represented as the kappa value) between ELISA, DENCHIK and commercially available q RT-PCR kits for DENV and CHIKV detection, were calculated using the equation below

| Reference Standard |  |  |  |
| --- | --- | --- | --- |
|  | Disease present | Disease absent | Total |
| Index Test positive | True positive (TP) | False positive (FP) | TP + FP |
| Index Test negative | False negative (FN) | True negative (TN) | TN + FN |
| Total | TP + FN | TN + FP |  |

Sensitivity = $TP / (TP + FN)$
Specificity = $TN / (TN + FP)$
Positive predictive value (PPV) = $TP / (TP + FP)$
Negative predictive value (NPV) = $TN / (TN + FN)$
Positive likelihood ratio (LR+) = $sensitivity / (1 - specificity)$
Negative likelihood ratio (LR-) = $(1 - sensitivity) / specificity$ .

The interrater agreement between the first and second interpreters for each assay (ELISA vs q RT-PCR) was assessed using Kappa statistics (k). Kappa agreement was calculated using *irr* package [28] Kappa is a measure of the degree of non-random agreement between observers or measurements of the same categorical variable. Agreement is considered as slight if the values are between 0.01-0.20; fair if kappa is between 0.41-0.60; good if kappa is between 0.60 and 0.80 and very good if greater than 0.80 [30,31].

Using Generalized Linear Model (GLM), we tested how DENV and CHIKV prevalence varies by month and detection methods. We modelled the infection status (infected or not infected, binomial distribution) as a response variable and interaction between detection method (IgM, NS1, RT-qPCR) and month as response variables[32].

## 3. Results

### 3.1 DENCHIK assay

DENCHIK assay showed simultaneous amplification of DENV serotypes and CHIKV and LoD of 10 viral copies per µl. (Fig 2A). (Supplementary Table 3)

**Fig 2:**
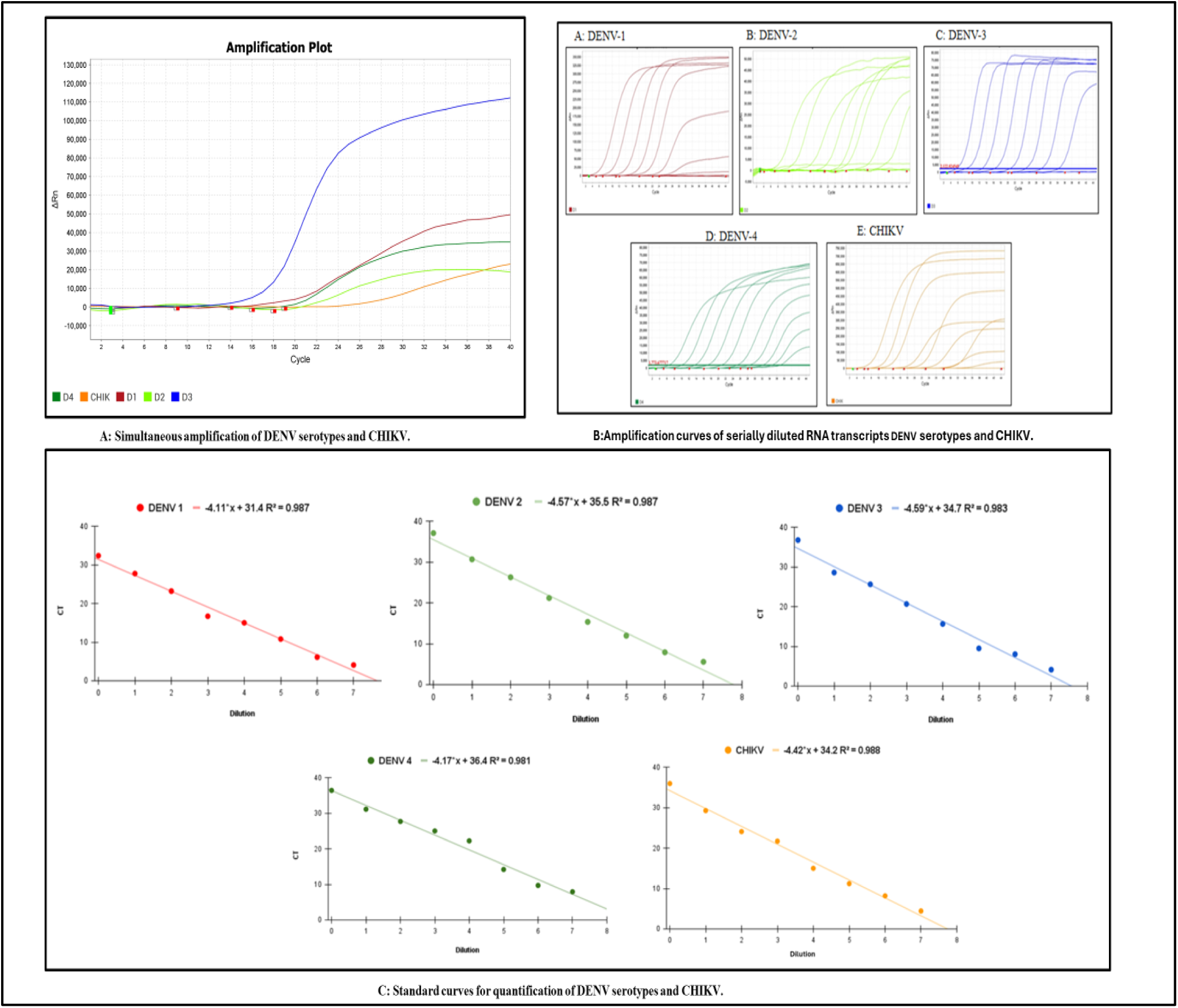
A: DENCHIK assay for detection of DENV and CHIKV simultaneously; B: Amplification plots depicting limit of detection (LoD) through serially diluted invitro RNA transcripts of both DENV and CHIKV; C: Calculation of LoD (viral copies/µl) DENV and CHIKV by plotting standard curves.

### 3.2 Estimation of DENV and CHIKV prevalence using DENCHIK and ELISA

The workflow for detection of DENV and CHIKV using ELISA and PCR is provided in Fig.3 and corresponding metadata is provided in Supplementary Table 1.

**Fig 3:**
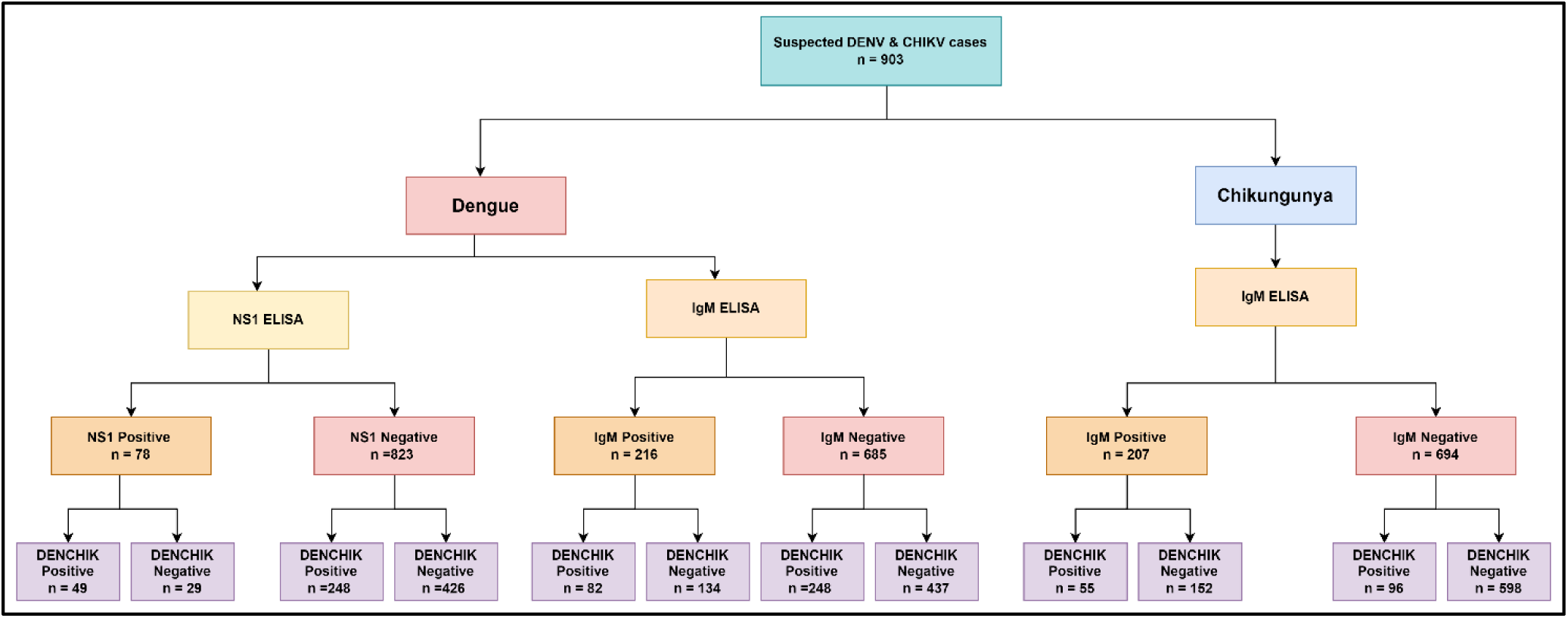
Flowchart of detection of DENV and CHIKV from clinical samples.

Using DENCHIK assay, the DENV prevalence 36.54% was significantly higher compared to 9% using NS1 ELISA (χ^2^=20.58, p<0.0001). However, DENCHIK showed no significant difference (23.9%) using IgM (χ^2^=1.54, p=0.0641). Similarly, DENCHIK assay (17%) showed no significant difference in CHIKV infections with IgM (23%) (χ^2^=1.125, p=0.28). We found similar pattern in detection of co-infections with DENV (14%) and CHIKV (8%) using both NS1 and IgM ELISA respectively, for detection Dengue and Chikungunya, and DENCHIK assay (χ^2^=1.83, p=0.17) (Fig 4).

**Fig 4:**
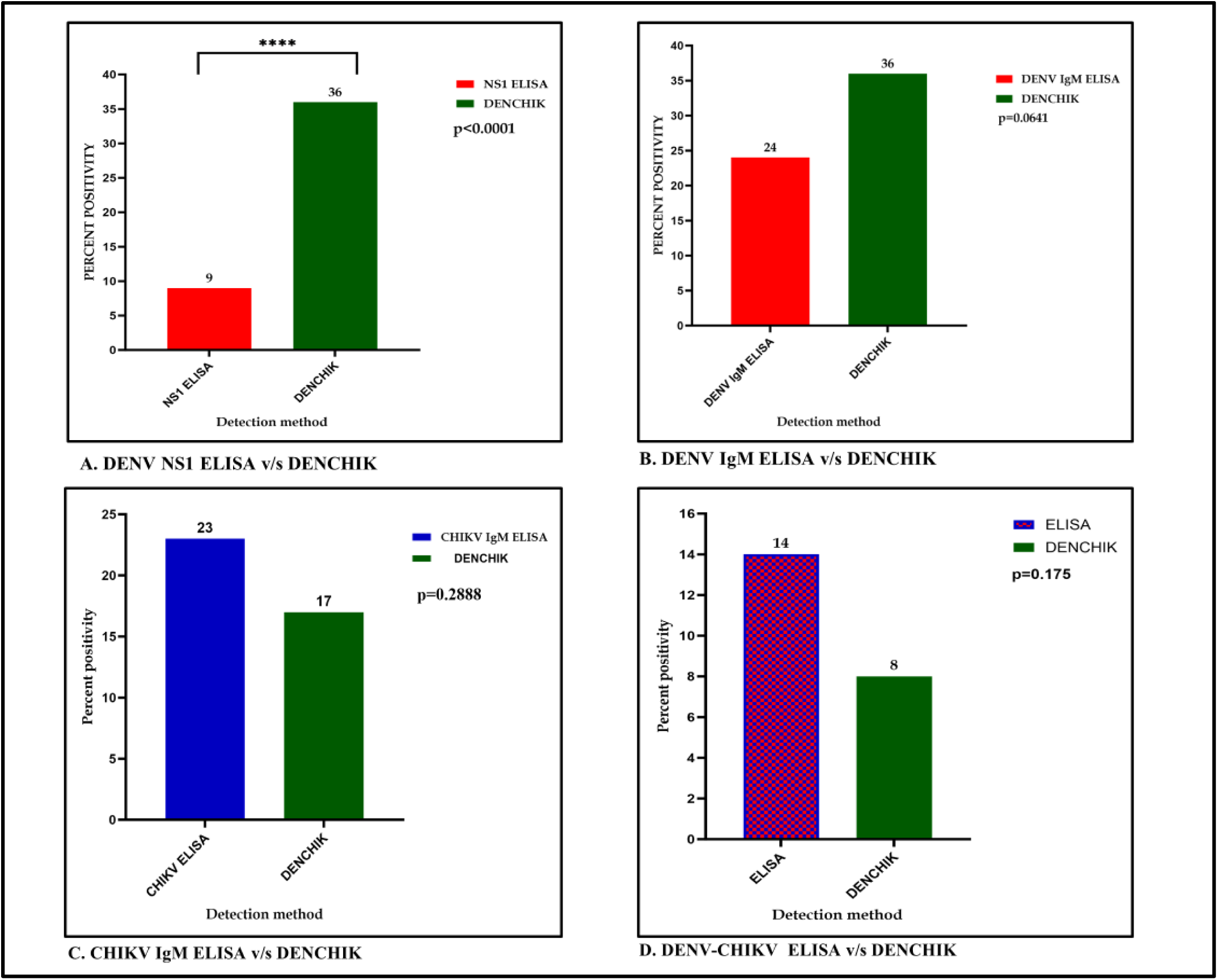
Comparison of prevalence of Dengue, Chikungunya and Co-infection through ELISA and DENCHIK assay.

DENCHIK assays detected all the four DENV serotypes. Overall, 36% of samples were infected with DENV. CHIKV was detected in 151 samples (17 %), whereas DENV-CHIKV co-infections were found in 76 clinical samples (8%). There was no significant difference observed in prevalence of DENV and CHIKV amongst the screened clinical samples (χ^2^=0.02 p=0.992) (Fig. 5).

**Fig. 5:**
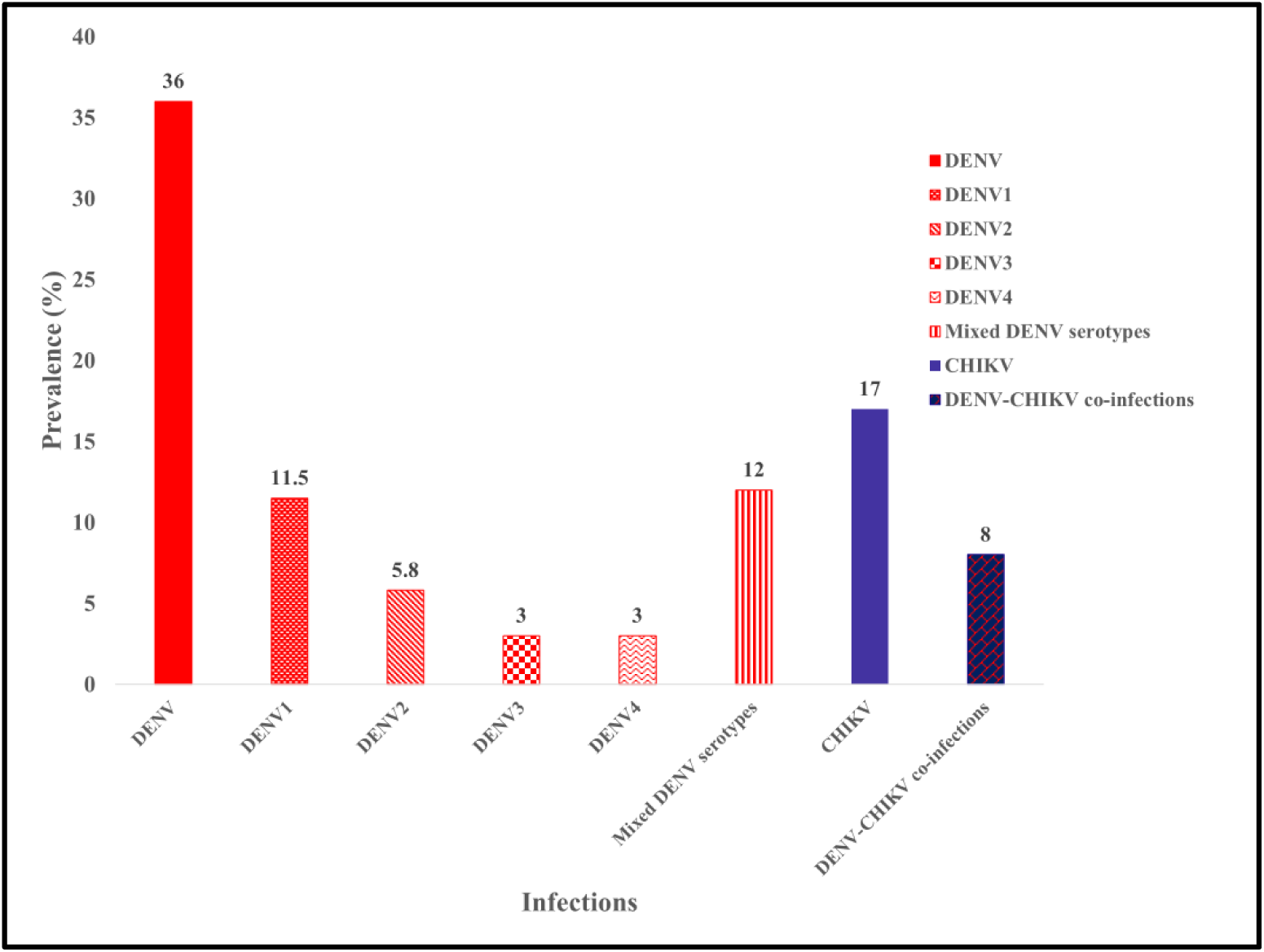
Prevalence of DENV serotypes, CHIKV and DENV-CHIKV co-infections in clinical samples.

We found, DENV1 (D1) serotype as the most prevalent serotype (104/903;11.5%) followed by DENV2 (D2) (54/903; 5.8%), DENV3 (D3) (32/903;3.5%) and DENV4 (D4) (29/903;3.2%). Mixed DENV serotype infections with D2, D3 and D4 serotypes were observed in 112 samples (12.4 %).

The geospatial distribution of DENV and, CHIKV infections appeared to be uniform across the city. (Fig 6).

**Fig 6:**
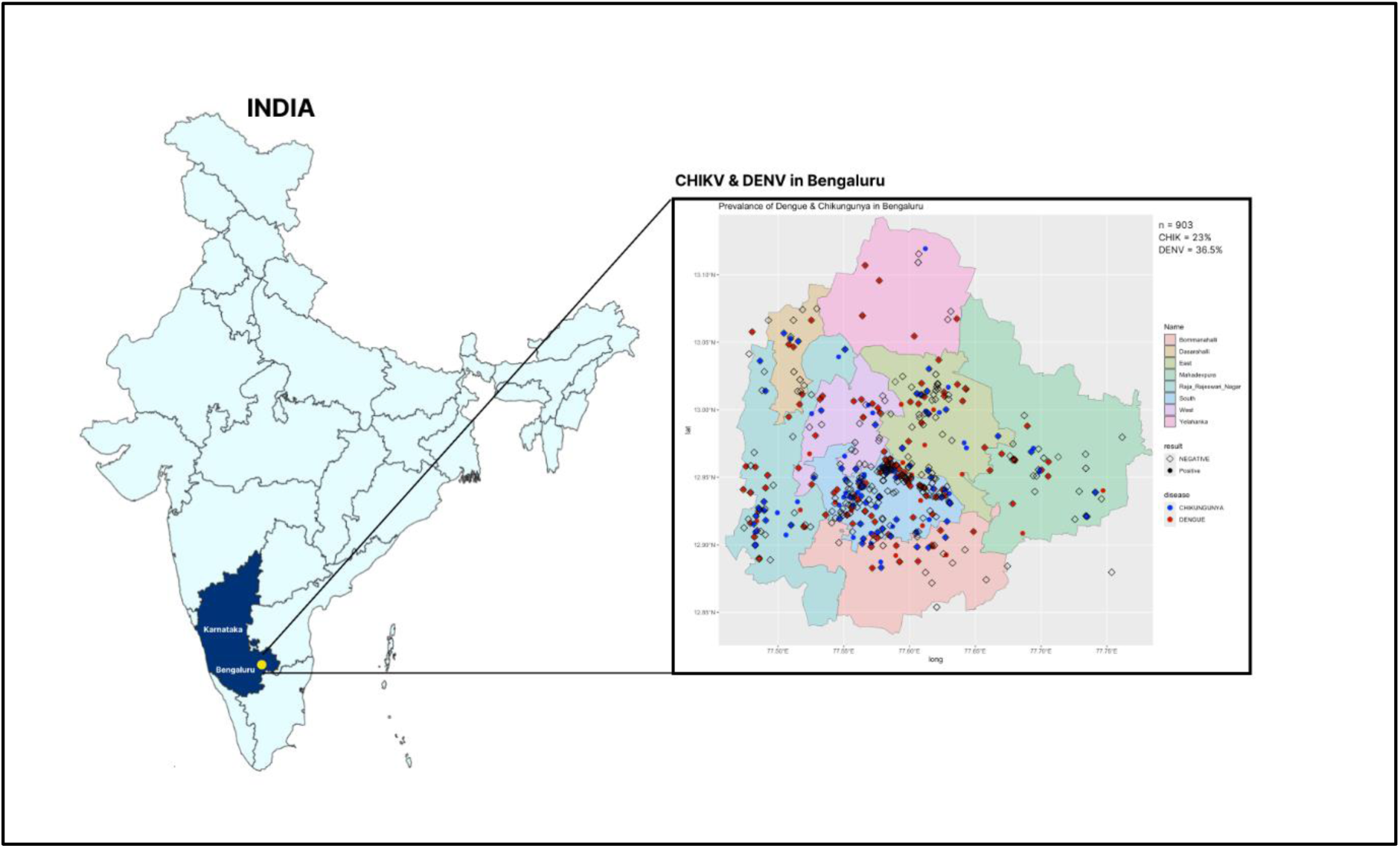
Mapping of prevalence of DENV and CHIKV cases in Bengaluru city.

We classified prevalence by age, gender and symptoms. Amongst the clinical samples that presented acute febrile illnesses, (n=555), fever (89%) myalgia (33%) and headache (33%) were found to be the most frequently reported symptoms (Table 2).

In comparison to ELISA, DENCHIK assay exhibited increased detection of DENV infection across months and age groups (Fig. 9). However, in case of CHIKV infections, IgM ELISA exhibited increased detection across the months (Fig. 10). Both DENCHIK and ELISA showed a high prevalence in July to September in Dengue and Chikungunya and low prevalence in October to December (Fig. 9 and Fig. 10).

**Fig 9:**
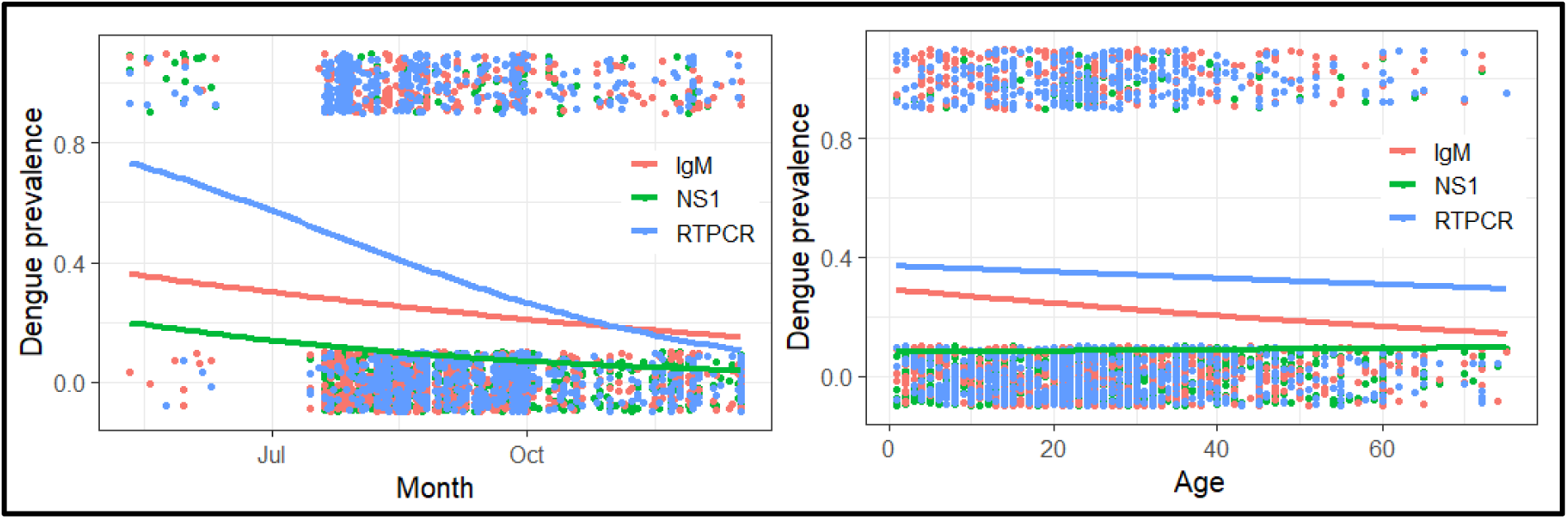
Detection of DENV and CHIKV by NS1 & IgM ELISA and DENCHIK (qRTPCR) across months and age.

**Fig 10:**
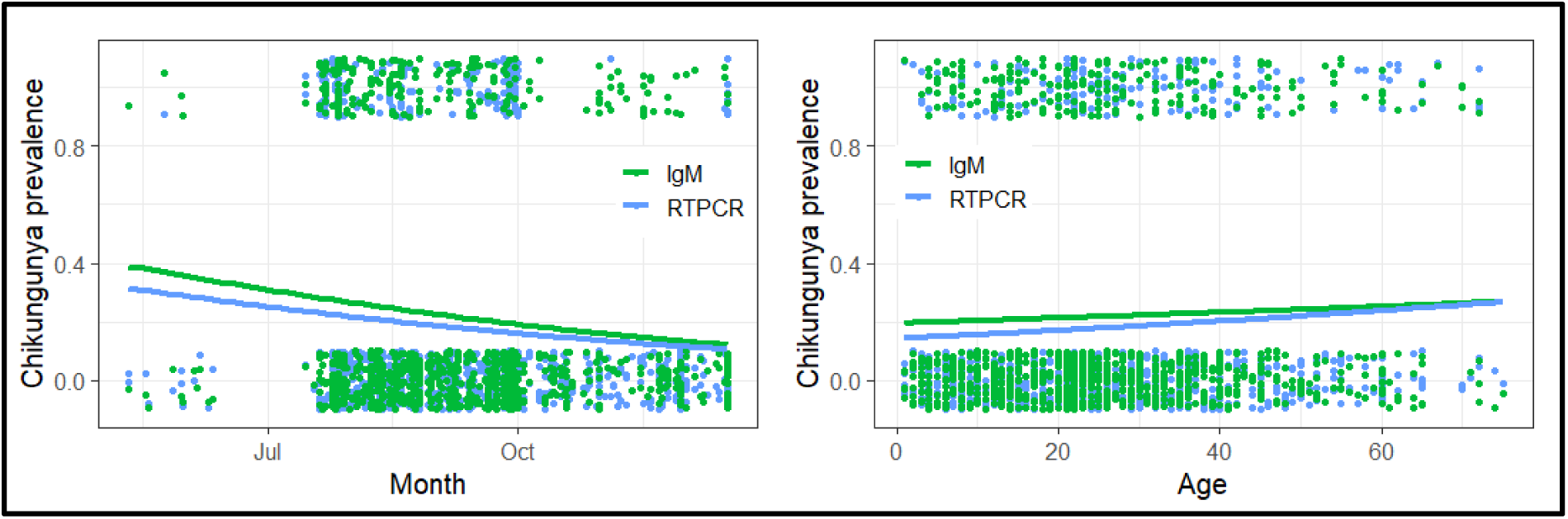
Detection of CHIKV by IgM ELISA and DENCHIK (RTPCR) across months and age

GLM showed that ELISA (NS1) showed negative association with Dengue detection whereas RTPCR showed marginally positive association. the interactions between detection methods and month were the best model predicting positive association in Dengue detection in July using q RT-PCR whereas NS1 test in June. The Dengue detection in general was lowest in September with a positive association with NS1 (Fig. 11).

**Fig 11:**
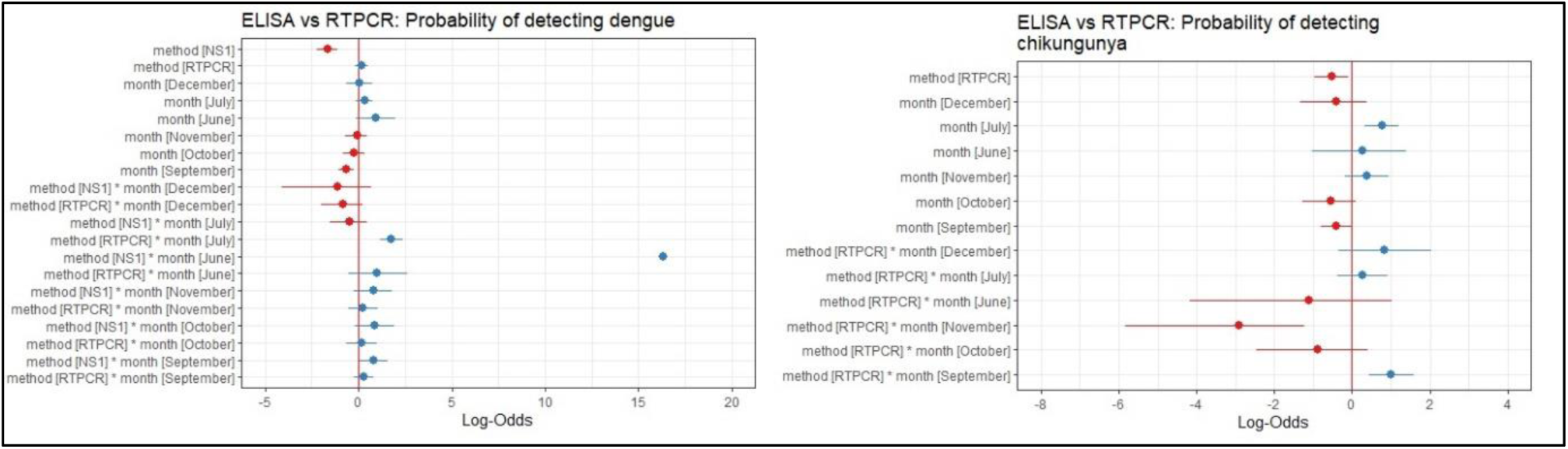
Interactive models of detection methods for Dengue and Chikungunya across months.

For Chikungunya, qRT PCR showed significant negative association. However, the interactive model was the best in predicting high infections in September with a positive interaction with q RT-PCR and significant negative association in November (Fig 11).

The patient age group did not affect the prevalence of DENV and CHIKV across the clinical samples screened using DENCHIK (Fig 9,10).

### 3.4 Comparison of diagnostic accuracies of ELISA,DENCHIK and commercial kits

#### 3.4.1 Diagnostic accuracy of NS1 & IgM assays with reference to DENCHIK for DENV and CHIKV detection

The sensitivity and specificity of DENCHIK assay in comparison to NS1 ELISA and IgM ELISA was 15% and 95% for DENV detection (Table 3). Similarly, the IgM ELISA depicted a sensitivity and specificity of 24% and 76%, respectively. IgM ELISA was observed to be 36.42% sensitive and 80% specific for CHIKV detection.

**Table 3:** Comparison of sensitivity, specificity, and agreement of ELISA, in comparison to DENCHIK.

| Index test/<br>Parameter<br>tested | Reference<br>standard | Infection<br>detected | Sensitivity<br>(%) | Specificity<br>(%) | Positive<br>predictive<br>value<br>(PPV)<br>% (95%<br>CI) | Negative<br>predictive<br>value<br>(NPV), %<br>(95% CI) | Likeliho<br>od Ratio<br>(LR)<br>(95%CI) | Odds Ratio<br>(DOR)<br>(95% CI) | Relative<br>Risk<br>(RR) (95%<br>CI) | Attributable<br>Risk<br>(AR)<br>(95% CI) |
| --- | --- | --- | --- | --- | --- | --- | --- | --- | --- | --- |
| NS1 ELISA | DENCHIK | DENV | 0.15<br>(0.11-0.20) | 0.95<br>(0.93-0.97) | 0.69<br>(0.55-0.79) | 0.64<br>(0.59-0.67) | 3.41 | 3.83<br>(2.09-6.80) | 1.89<br>(1.49-2.29) | 0.32<br>(0.17-0.44) |
| IgM ELISA | DENCHIK | DENV | 0.24<br>(0.20-0.29) | 0.76<br>(0.73-0.80) | 0.38<br>(0.31-0.44) | 0.64<br>(0.60-0.67) | 1.05 | 1.06<br>(0.77-1.46) | 1.03<br>(0.84-1.25) | 0.013<br>(-0.06-0.09) |
| IgM ELISA | DENCHIK | CHIKV | 0.36<br>(0.29-0.44) | 0.80<br>(0.76-0.82) | 0.27<br>(0.21-0.33) | 0.86<br>(0.83-0.88) | 1.80 | 2.25<br>(1.53-2.38) | 1.921<br>(1.42-2.56) | 0.13<br>(0.06-0.20) |
| Dengue NS1<br>ELISA<br>symptoms<br>($< 5$ days) | DENCHIK | DENV | 0.20<br>(0.14-0.29) | 0.93<br>(0.89-0.95) | 0.53<br>(0.39-0.68) | 0.76<br>(0.70-0.81) | 3.07 | 3.61<br>(1.87-7.20) | 2.20<br>(1.51-3.03) | 0.29<br>(0.12-0.46) |
| Dengue NS1<br>ELISA<br>symptoms<br>($> 5$ days) | DENCHIK | DENV | 0.19<br>(0.10-0.34) | 0.97<br>(0.90-0.99) | 0.80<br>(0.49-0.96) | 0.67<br>(0.58-0.76) | 6.82 | 8.24<br>(1.80-39.53) | 2.44<br>(1.41-3.53) | 0.47<br>(0.10-0.66) |
| Dengue IgM<br>ELISA<br>symptoms<br>($< 5$ days) | DENCHIK | DENV | 0.23<br>(0.13-0.38) | 0.74<br>(0.65-0.81) | 0.26<br>(0.15-0.42) | 0.71<br>(0.62-0.80) | 0.90 | 0.88<br>(0.39-1.99) | 0.90<br>(0.49-1.6) | 0.02<br>(-0.13-0.21) |
| Dengue IgM<br>ELISA<br>symptoms<br>($> 5$ days) | DENCHIK | DENV | 0.30<br>(0.24-0.36) | 0.74<br>(0.69-0.80) | 0.43<br>(0.35-0.50) | 0.62<br>(0.58-0.67) | 1.17 | 1.24<br>(0.84-1.81) | 1.14<br>(0.90-1.41) | 0.50<br>(-0.04-0.15) |
| Chikungunya<br>symptoms<br>< 5 days | DENCHIK | CHIKV | 0.24<br>(0.17-0.33) | 0.77<br>(0.73-0.82) | 0.27<br>(0.19-0.36) | 0.76<br>(0.71-0.80) | 1.08 | 1.10<br>(0.60-1.86) | 1.08<br>(0.73-1.5) | 0.02<br>(-0.08-0.13) |
| Chikungunya<br>symptoms<br>> 5 days | DENCHIK | CHIKV | 0.28<br>(0.12-0.51) | 0.74<br>(0.66-0.80) | 0.12<br>(0.05-0.25) | 0.89<br>(0.82-0.93) | 1.06 | 1.08<br>(0.40-3.25) | 1.07<br>(0.41-0.27) | 0.07<br>(-0.09-0.16) |

NS1 ELISA, in comparison to DENCHIK exhibited a diagnostic sensitivity of 20% and 19% for detection of DENV in category of <5 days and >5 days post symptom presentation. However, specificity of NS1 ELISA in these both categories was found to be 93% and 97% respectively (Table 3).

Upon comparing diagnostic accuracy of IgM ELISA in comparison to DENCHIK, we observed detection sensitivity of ELISA in clinical samples presenting symptoms for <5 days and >5 days to be 23% and 30% for DENV, and 24% and 28% sensitive for CHIKV respectively. However, ELISA was found to be 74% specific for DENV and 77% and 74 % for CHIKV in both <5 and >5 days categories of symptom presentation (Table 3).

The detection sensitivity of DENCHIK compared with NS1 antigen ELISA and IgM ELISA as reference. DENCHIK exhibited sensitivity and specificity of 65% and 63% and 37.96 and 66.45%, respectively. DENCHIK was found to be 26.57% sensitive and 86.07 % specific for CHIKV detection as compared to IgM ELISA. Similarly, DENCHIK was evaluated for detection of DENV and CHIKV in the dataset according to the days of onset of symptoms (Table 4).

**Table 4:** Comparison of sensitivity, specificity, and agreement of DENCHIK, in comparison to ELISA.

| Index test/<br>Parameter<br>tested | Reference<br>standard | Infection<br>detected | Sensitivity | Specificity | Positive<br>predictive<br>value<br>(PPV)<br>(95% CI) | Negative<br>predictive<br>value<br>(NPV), (95%<br>CI) | Likeliho<br>od Ratio<br>(LR) | Odds Ratio<br>(DOR)<br>(95% CI) | Relative<br>Risk<br>(RR) (95%<br>CI) | Attributabl<br>e Risk<br>(AR)<br>(95% CI) |
| --- | --- | --- | --- | --- | --- | --- | --- | --- | --- | --- |
| DENCHIK | NS1<br>ELISA | DENV | 0.65<br>(0.54-0.74) | 0.63<br>(0.59-0.66) | 0.18<br>(0.14-0.23) | 0.94<br>(0.91-0.95) | 1.80 | 3.26<br>(2.01-5.24) | 2.84<br>(1.86-4.34) | 0.11<br>(0.06-0.17) |
| DENCHIK | IgM<br>ELISA | DENV | 0.38<br>(0.31-0.44) | 0.63<br>(0.6-0.67) | 0.24<br>(0.20-0.29) | 0.77<br>(0.72-0.80) | 1.03 | 1.06<br>(0.71-1.46) | 1.04<br>(0.82-1.32) | 0.01<br>(-0.04-0.07) |
| DENCHIK | IgM<br>ELISA | CHIKV | 0.26<br>(0.21-0.32) | 0.86<br>(0.83-0.88) | 0.36<br>(0.29-0.44) | 0.79<br>(0.76-0.82) | 1.92 | 2.25<br>(1.53-3.29) | 1.79<br>(1.38-2.29) | 0.16<br>(0.08-0.25) |
| DENCHIK | Dengue<br>NS1<br>ELISA<br>symptoms<br>($< 5$ days) | DENV | 0.31<br>(0.20-0.44) | 0.76<br>(0.70-0.80) | 0.20<br>(0.12-0.29) | 0.85<br>(0.80-0.89) | 1.28 | 1.40<br>(0.74-2.65) | 1.32<br>(0.78-2.18) | 0.04<br>(-0.04-0.15) |
| DENCHIK | Dengue<br>NS1<br>ELISA<br>symptoms<br>($> 5$ days) | DENV | 0.85<br>(0.67-0.94) | 0.57<br>(0.51-0.63) | 0.17<br>(0.12-0.25) | 0.97<br>(0.93-0.98) | 2.00 | 7.79<br>(2.753-21.30) | 6.59<br>(2.45-8.65) | 0.15<br>(0.07-0.23) |
| DENCHIK | Dengue<br>IgM<br>ELISA<br>symptoms | DENV | 0.62<br>(0.44-0.77) | 0.74<br>(0.68-0.79) | 0.20<br>(0.13-0.30) | 0.95<br>(0.90-0.97) | 2.37 | 4.60<br>(2.07-9.92) | 3.07<br>(1.93-7.73) | 0.15<br>(0.06-0.26) |
|  | (< 5 days) |  |  |  |  |  |  |  |  |  |
| DENCHIK | Dengue<br>IgM<br>ELISA<br>symptoms<br>(> 5 days) | DENV | 0.74<br>(0.56-0.81) | 0.86<br>(0.61-0.85) | 0.26<br>(0.12-0.35) | 0.92<br>(0.81-0.96) | 2.17 | 3.85<br>(1.98-9.12) | 2.97<br>(1.71-8.67) | 0.14<br>(0.05-0.29) |
| DENCHIK | Chikungu<br>nya IgM<br>symptoms<br>(< 5 days) | CHIKV | 0.16<br>(0.1-0.25) | 0.90<br>(0.87-0.92) | 0.27<br>(0.17-0.40) | 0.81<br>(0.77-0.85) | 1.51 | 1.60<br>(0.83-3.14) | 1.44<br>(0.86-2.27) | 0.08<br>(-0.03-0.23) |
| DENCHIK | Chikungu<br>nya IgM<br>symptoms<br>(> 5 days) | CHIKV | 0.35<br>(0.27-0.44) | 0.82<br>(0.77-0.86) | 0.41<br>(0.32-0.51) | 0.77<br>(0.72-0.81) | 1.93 | 2.43<br>(1.50-3.90) | 1.84<br>(1.34-2.49) | 0.19<br>(0.08-0.30) |

DENCHIK, in comparison to NS1 ELISA exhibited sensitivity of 31% and 85% for detection of DENV in category of <5 days and >5 days post symptom presentation. However, specificity of DENCHIK in these both categories was found to be 76% and 57% respectively (Table 4).

On comparing the detection sensitivity of DENCHIK with respect to IgM ELISA, we found sensitivity of 65% and 74% for DENV detection and 16% and 35% for CHIKV in both <5 days and >5 days post symptom presentation categories. The specificity of DENCHIK was recorded to be 74% and 86% for DENV and 90% and 82% for CHIKV, respectively (Table 4).

For comparison between ELISA and DENCHIK, both interpreters show slight agreement with kappa of 0.121 (DENCHIK vs NS1) and 0.109 (DENCHIK vs IgM) in DENV detection and kappa of 0.140 for CHIKV detection using DENCHIK vs IgM.

#### 3.4.2 DENCHIK versus commercial kits

We used a subset of samples (n=100) to compare DENCHIK with commercially available and widely used RT-PCR kits (Altona Diagnostics, Hamburg, Germany) and Real Star Chikungunya RT-PCR kit (Altona Diagnostics, Hamburg, Germany), for detection of DENV serotypes and CHIKV respectively. We observed 100% concordance in detection of DENV, DENV serotypes and CHIKV across DENCHIK and respective commercial kits. DENCHIK exhibited >95%sensitive and specificity for the detection of DENV, DENV serotypes and CHIKV. (Table 6A).

**Table 6A:**
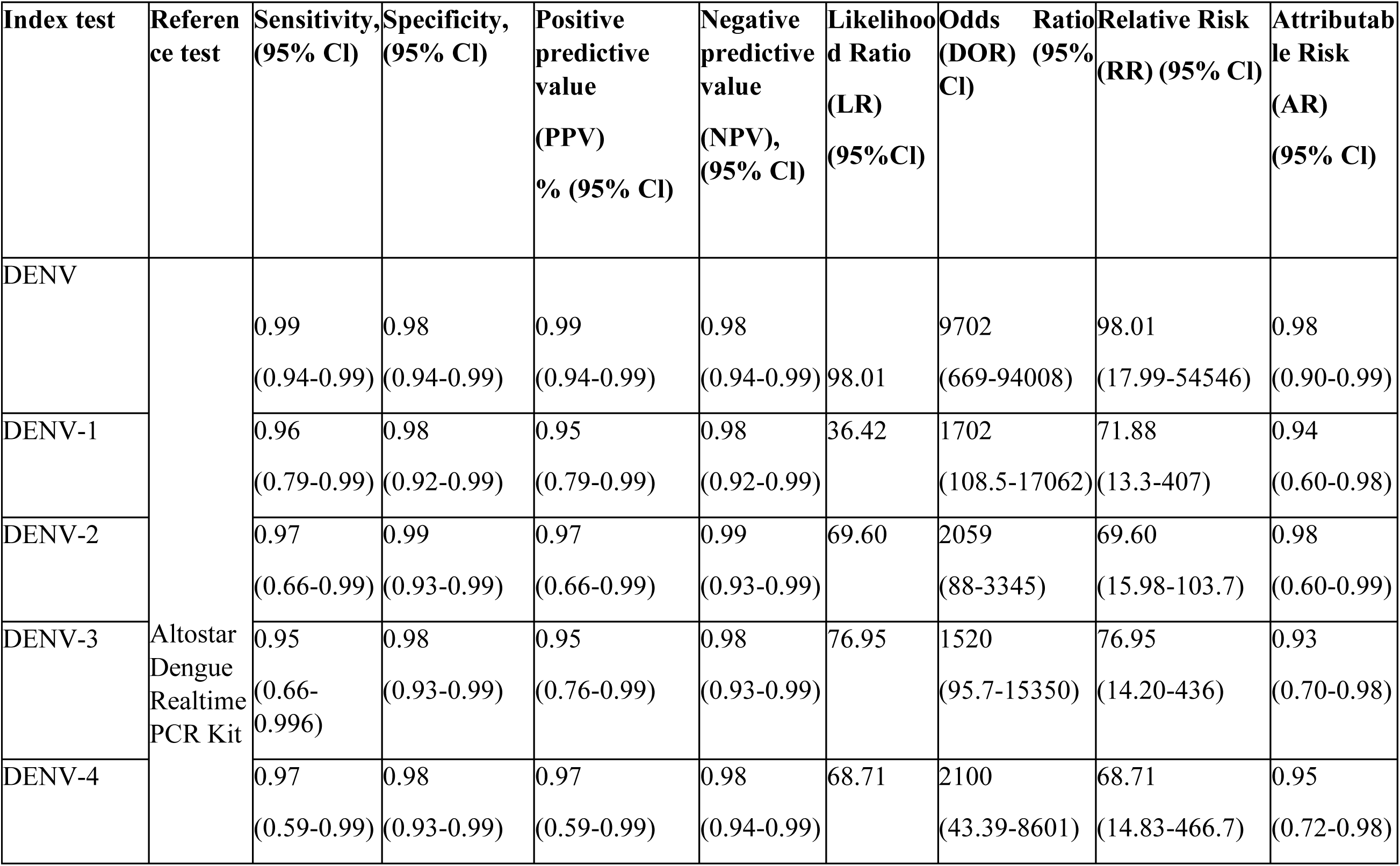
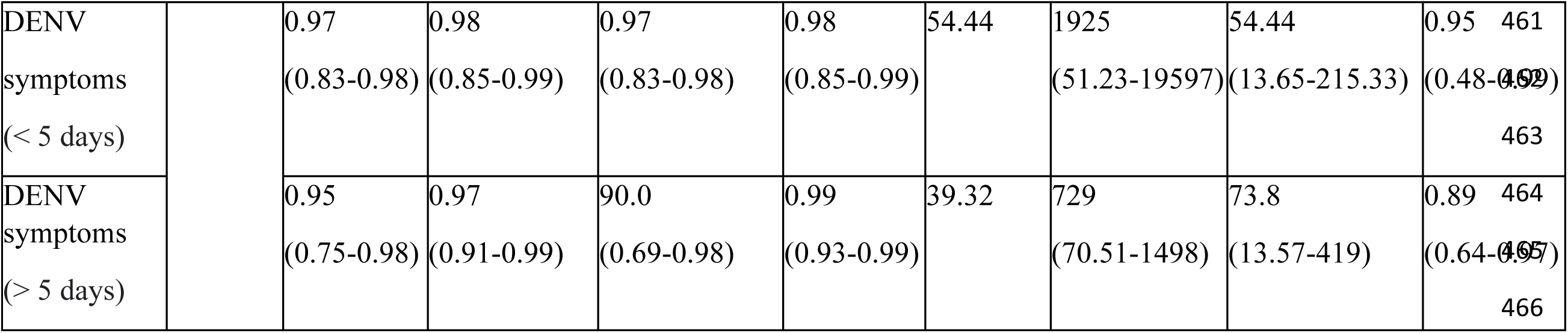
Diagnostic accuracy of DENCHIK for DENV serotype detection w.r.t commercially available RT-PCR Kits.

DENCHIK could detect DENV and CHIKV with 99% and 98% sensitivity and 98% specificity respectively, upon comparison to commercially available kits (Table 6A, 6B). Further, DENV serotypes DENV1,2,3 and 4 were 96%,97%,95%and 97% sensitive and demonstrated 98%,99%,98% and 98% specificity respectively.

**Table 6B:**
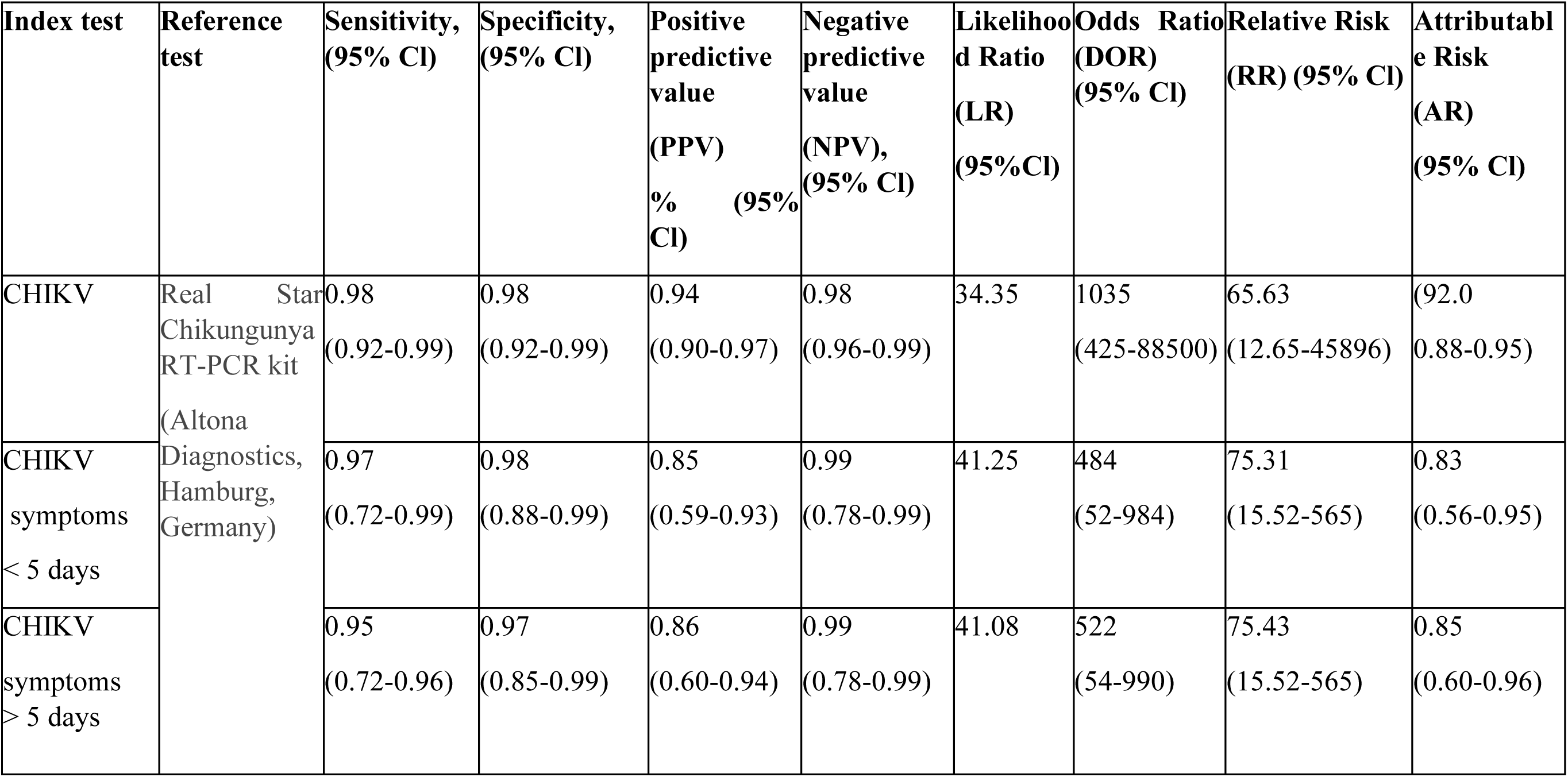
Diagnostic accuracy of DENCHIK for CHIKV detection w.r.t commercially available RT-PCR Kits.

From a subset of 555 clinical samples that presented acute febrile illnesses and various symptoms, DENV detection >5 days and <5 days post symptom presentation was observed to be 97% and 95% sensitive and with 98% and 97% specificity respectively. However, in case of CHIKV detection >5 days and <5 days post symptom presentation exhibited 97% and 95% sensitivity and 98% and 97% specificity, respectively.

## 4. Discussion

Early detection of DENV and CHIKV is crucial, with major public health efforts being concerted towards accurate diagnostics, accelerated surveillance, and vector control for prompt mitigation of the disease. NS1 antigen ELISA and rapid tests, are routinely performed for early diagnosis of DENV infections. However, ELISA remains the only widely used assay for diagnosing CHIKV infections in clinical samples [33].There is a lack of accurate nucleic acid-based diagnostic assays for early, rapid and accurate detection of DENV and CHIKV in tropical countries..

We developed a multiplex q RT-PCR assay, DENCHIK which is highly sensitive and specific, for simultaneous detection and quantification of all four serotypes of DENV and CHIKV. Currently, there are no such assays being used in health care system for systematic monitoring of Dengue and Chikungunya burden in India.

DENCHIK demonstrated a reasonable limit of detection (LoD), of approximately ten viral copies per microlitre for DENV and CHIKV implying that the assay is sensitive for detection of low viral titres in clinical samples. Furthermore, serotype-based DENV detection revealed 8.5 % of mixed infection with DENV as well as CHIKV. Previous reports emphasize the need for timely diagnosis Of DENV-CHIKV co-infections to prevent multi-organ dysfunction, leading to higher mortality than mono infections [34,35].

DENCHIK detected 7.6% higher number of DENV infections in comparison to ELISA . However, in case of CHIKV, IgM ELISA detected 6.65% more infections in comparison to DENCHIK. This might be due to the questionable sensitivity and specificity of IgM ELISA assay for Chikungunya diagnosis. A study from Manipal, in Karnataka, India have reported prolonged persistence of antibodies, until 10 months after CHIKV infection [36]. These further questions the reliability of IgM ELISA assay to detect acute CHIKV infection and presence of virus based solely on detection of antibodies.

Higher detection and prevalence of DENV and CHIKV infections by both ELISA and DENCHIK was observed across June to September, coinciding with the peak monsoon season in Bengaluru. This observation aligns with the findings of Dharmamuthuraja et al ,wherein a similar pattern for *Aedes* mosquito larval habitat was observed coinciding with peak in Dengue incidence in Bengaluru [4].However DENCHIK could as well detect higher prevalence of DENV infections in months from October to December highlighting the sensitivity of the assay.

Using DENCHIK assay, we recorded 36% of DENV infections,17% of CHIKV and 8 % of DENV-CHIKV co-infections, in the clinical samples, which were otherwise underestimated or overestimated by only ELISA-based tests respectively.

Furthermore, we report high prevalence and co-circulation of all four DENV serotypes in Bengaluru. The persistence and prevalence of all four DENV serotypes could lead to immunity-driven co-evolution of Dengue virus strains and hence it is crucial to know the prevalence and distribution of DENV serotypes to understand the evolution of the Dengue virus and predict future outbreaks. In a recent report, Jagtap et al, analysed 119 dengue genomes across India to understand the complexity of dengue virus evolution [37]. However, DENCHIK is cost effective way to understand spatio-temporal distribution of serotype prevalence where only a subset of samples can be sequenced to understand viral evolution.

While there are reports of serotype-based outbreaks from several states in India and Nepal, DENV1.DENV-3 and DENV-4 outbreaks have been observed and reports across India in the states of Karnataka, Agra, Odisha and West Bengal, using serology, molecular typing and RT-PCR based assays [38–43],however the DENV serotype landscape still remains to be further explored in metropolitan cities of India, such as Bengaluru, where Dengue is a huge public health concern.

Amongst the mixed serotype infections, D1:D2 were the most prevalent mixed serotype infection observed. Both these serotypes have been associated with major outbreaks in Nepal [44]and Mexico [45] ,mixed serotype and the circulation also suggests the higher endemicity of both of these serotypes or might as well be a result of the viral transmission through *Aedes* mosquito.[32,41,46].

Molecular surveillance of Dengue, using DENCHIK could be used as an complimentary cost-effective tool in parallel to the whole genome sequencing approach to understand the persistence and prevalence of multiple DENV serotypes in clinical samples contributing to increase in disease endemicity. Also, DENCHIK can be employed to understand the temporal and spatial prevalence of DENV serotypes across the city and further aid in informed intervention in areas with changing serotype transmission dynamics.

DENV and CHIKV prevalence showed no significant difference by age groups and genders. However, fever and myalgia, were the two prominent symptoms observed across all the clinical samples. This observation is concordant with earlier report, wherein symptoms as such, are common across various other infections such as flu, malaria, and other bacterial and viral illnesses, making targeted molecular surveillance imperative for prevention and control of priority arboviral illnesses [47].

On comparing the detection accuracy of DENCHIK with ELISA and vice-versa using both as reference standards against the other respectively, we inferred DENCHIK to be more sensitive than specific for detection of DENV. However, ELISA shows better specificity, but lower sensitivity for Dengue diagnosis. This observation coincides with a previous finding [48],wherein ELISA displays lower sensitivity as compared to nucleic acid based diagnostic assays [48].

However, in case of CHIKV infections, we found ELISA to be more sensitive and DENCHIK to be more specific for detection of Chikungunya.

However, upon comparing DENCHIK with commercially available RT-PCR, serotyping kits, DENCHIK was found to >98% sensitive and specific for detection of all four DENV serotypes and CHIKV respectively, suggesting DENCHIK to be more reliable molecular assay for accurate disease diagnostics.

DENCHIK also enabled detection of DENV-CHIKV coinfection with 96% sensitivity and 98% specificity. The diagnostic accuracies calculated across the four serotypes revealed more than 90% sensitivity and specificity of the assay.

DENCHIK demonstrated increased sensitivity and specificity, across clinical samples presenting symptoms less than 5 days as well as greater than 5 days post onset of both DENV and CHIKV infections. DENCHIK thus outperforms both NS1 and IgM ELISA tests respectively, as they demonstrate preferred and conditional accuracy for disease diagnosis, in either less than 5 days or more than 5 days post symptom presentation, clinical scenarios.[9,50].These findings justify the potential of DENCHIK for rapid, reliable, and accurate detection method for DENV, CHIKV and DENV-CHIKV co-infections, in clinical settings.

Molecular assays, although preferable and accurate are often ignored owing to the costs involved in consumables and dependence on technical expertise. Hence, cost effective molecular assays, employed across primary health centres as well as tertiary care hospitals would aid early and exact determination of priority arboviral infections such as DENV and CHIKV across the world.

Molecular surveillance using DENCHIK will enhance our understanding of the serostatus of a DENV infected individual, as well as the probable genetic variations, in the virus, which in turn might affect the infection dynamics in an endemic situation [51–53]. In summary the introduction of molecular detection of priority infectious diseases is the way forward for rapid and accurate estimation of the disease burden in the ecosystem.

## 5. Conclusions

The study was proposed to understand the presence of arboviral diseases such as Dengue Chikungunya and co-infections in patients presenting acute febrile illnesses. PCR-based diagnostics have gained momentum in the recent past for the detection of the SARS-CoV2 virus infections during the global pandemic, however arboviral diseases, long affecting human health are still being diagnosed by ELISA. This has often led to inaccurate estimation of infection prevalence and disease burden in urban Bengaluru. Our findings suggest the efficiency of DENCHIK, an inhouse multiplex q RT-PCR assay, developed for screening DENV, CHIKV and Co-infections in clinical samples presenting acute febrile illnesses cases, from as early as day 2 of symptom presentation.

The simultaneous five-plex detection of DENV serotypes and CHIKV developed in this study led to specific and sensitive detection of DENV, CHIKV, and co-infections. The results demonstrate accurate, sensitive, and specific detection of these arboviral infections as compared to routine serology.

The study intends to propose systematic diagnostic approach in public health centres to screen acute febrile illnesses for detection of priority vector-borne diseases through advanced molecular methods, allowing accurate and timely detection, treatment, prevention, and control of the spread of arboviral infections in low resource settings.

## Supporting information

https://tatainstitute-my.sharepoint.com/:x:/g/personal/mansi_malik_tigs_res_in/EcV2MpS7DcRDodgQE-EhW7sBpBvuMsOamz2Pj2qh65yGRw?e=zBArO8

## Data Availability

All the data pertaining to the manuscript findings has been included with the submission.

## 6. Acknowledgements

This study is financially supported by Tata Trusts funding to Tata Institute for Genetics and Society. We acknowledge ICMR-NIV for providing Chikungunya IgM ELISA kits. We sincerely acknowledge the administrative and the hospital staff of Bruhat Bengaluru Mahanagar Palike, Bengaluru, and Bangalore Medical College and Research Centre (BMCRI) for their support in the successful execution of this study.

## 7. Author declaration

The DENCHIK assay has been filed for a Provisional Patent, vide application number, 202341070795, entitled, ‘MULTIPLEX QUANTITATIVE RTPCR ASSAY FOR DETECTION OF DENGUE AND CHIKUNGUNYA VIRUSES.’

## 8. Authors’ Contributions

Conceptualization: MRM, SU, FI & TC

Experimental design: SU, MM,

Lab experiments: MM, SW, RM, DSP, MAT

Data analysis: MM &FI

Project administration: MM and SU

Supervision: MM, SU, FI, TC & RKM Writing -Original draft: MM

Writing-review & editing: MM, FI, and SU

Resources and administration: RKM

